# Misinformation on covid-19 origin and its relationship with perception and knowledge about social distancing: A cross-sectional study

**DOI:** 10.1101/2020.10.06.20207894

**Authors:** Lenisse M. Reyes, Lilibeth Ortiz, Maxwell Abedi, Yenifel Luciano, Wilma Ramos, Pablo J. de Js. Reyes

**Affiliations:** M.D. Centro de Investigaciones Biomédicas Y Clínicas (CINBIOCLI), Department of Clinical Research, Hospital Regional Universitario José María Cabral Y Báez (HRUJMCB), Santiago, Dominican Republic; M.D. Independent researcher; M.Sc. Department of Forensic Sciences, University of Cape Coast, Cape Coast, Ghana; M.D. Department of Surgery, Hospital Metropolitano de Santiago (HOMS), Santiago, Dominican Republic

## Abstract

Despite the vast scientific evidence obtained from the genomic sequencing of COVID-19, a controversy regarding its origin has been created in the mass media. This could potentially have a long-term influence on the behavior among individuals, such as failure to comply with proposed social distancing measures, leading to a consequent rise in the morbidity and mortality rates from COVID-19 infection. Several studies have collected information about knowledge, attitudes and practices regarding COVID-19; however, very little is known about the relationship of the perceptions of the individuals regarding the origin of the virus with the knowledge and perception about social distancing. This study aimed at ascertaining this relationship. For such purpose, a web-based cross-sectional study was conducted among a sample population from five provinces of the Dominican Republic within the period of June to July of 2020. The data collection instrument exploited in the study was a self-designed questionnaire distributed throughout different social media platforms. A purposive sampling strategy was implemented and a total of 1195 respondents completed the questionnaire. The collected data was analyzed using SPSS. Descriptive statistics, stepwise multiple linear regression and one-way multivariate analysis were implemented to test the hypotheses. The level of education was significantly associated (*P* = 0.017) with individuals’ perception about the origin of COVID-19, whilst only age (*P* = 0.032) and education level (*P* < 0.001) statistically significantly predicted ‘knowledge about social distancing’. Perception of COVID-19 origin was statistically significant associated (*P* = < 0.001) with the measures of the dependent variables (knowledge and perception on social distancing). The present study has established a possible link between the ‘perception of COVID-19 origin’ and ‘the perception and knowledge about social distancing’.

## INTRODUCTION

Following the bubonic plague, there has been myriad well-recognized epidemics and pandemics worldwide [1,2], which have recorded a rapid increase in morbidity and mortality rates coupled with a disruption in the dynamics of environmental, ecological and socio-economic factors among humans [2,3]. Recent occurring pandemics are zoonotic in nature [3] due to the rapid growth rate among both the human and animal populations, thereby bridging the transmission gap between the two and consequently easing the expansion of such zoonotic infections globally [4,5]. In 2019, a brand-new viral infection emerged in Wuhan, China [6] caused by the coronavirus SARS-CoV-2, also known as COVID-19. The infection has been declared in the current year as a “Public Health Emergency of International Concern ‘‘ and has posteriorly become a pandemic [7].

The coronavirus family is characterized by a low fidelity RNA polymerase, nucleic acids with a high recombination frequency and an unusual extended genome, which facilitate their diversity and the emergence of viruses that can easily adapt to new hosts and environments [8-10]. The genomic sequence of SARS-CoV-2 has been documented to be akin to SARS-CoV [11]. Following the foregoing discovery, many other β-coronaviruses have been identified in both bats and humans. Notable among them was the BatCoV RaTG13 (isolated from *Rhinopulus affinis*), which shares about 96% of its genomic sequence with the novel SARS-CoV-2 [12,13]. Further, Pangolin-CoV harbored in Guangdong pangolins was again noted to have a genomic sequence very similar [12,14] to the amino acid residues of the receptor binding domain (RBD) of SARS-CoV-2 [13,15]. BatCoV RaTG13 [15], on the other hand, shares only one amino acid residue in the RBD as that of the novel coronavirus, despite sharing 96% homology otherwise. Moreover, a scientific theory published in a pre-printed repository vis-à-vis the origin of COVID-19 [16] found shared amino acids with those of the Human Immunodeficiency Virus (HIV-1) in the genomic sequence of SARS-CoV-2. The validity of such findings was immediately questioned and dismissed by several researchers, which subsequently led to a formal retraction by the authors and a withdrawal of the paper from the repository [17]. However, despite the withdrawal of the article and the reassurance made by other scientists who verified the genomic sequence of SARS-CoV-2 and the natural origin of the virus [18,19] these findings spread in the news and social media around the world, generating more controversy and reinforcing existing unofficial and popularly disseminated theories that established that the virus “leaked from a laboratory in Wuhan” and is probably a product of genetic manipulation in an effort to discover a vaccine for HIV-1[20-22].

In a short period of time, millions of infections and thousands of deaths have been reported from SARS-CoV-2 around the world. In the Dominican Republic, more than 100,000 cases have been recently reported with a case fatality ratio of 1.9% and a tendency to increase, similar to other countries of the region [23]. Owing to the ease of person-to-person transmission of SARS-CoV-2, a non-pharmacological intervention deemed ‘social distancing’ is currently being practiced to minimize spread of the virus [24]. Conventionally, early and sustained imposition of these interventions have been demonstrated to reduce mortality rates and flatten the epidemiologic curve significantly among varied countries, such as the United States, during the last registered pandemic in 1918 [25-26]. Currently, early interventions like social distancing have significantly limited the effects and slowed the transmission of COVID-19 in its stage of epidemic in mainland China [27] and New Zealand [28]. Among the Caribbean, the Dominican Republic has adapted the mandatory usage of nose masks in public places, as well as the suspension of small businesses, public transportation and a 10 hours’ curfew (7 p.m.-5 a.m.) as an additional measure to strengthen the social distancing protocols in an attempt to flatten the epidemiologic curve [29]. Despite these measures to contain the spread of the virus, some rule-breaking events have still occurred [30,31].

Various observational studies have been performed to assess the knowledge, attitudes and perceptions about COVID-19 among healthcare workers and the general populace, and some of them assess the perception and knowledge about the origin of COVID-19 and social distancing [32-35], yet very little literature exists on the relationship between an individual’s understanding of social distancing and their perceptions regarding the origin of SARS-CoV-2; it is possible that the controversy revolving around the origin of COVID-19 in the mass media is influencing the perceptions and knowledge about social distancing of the general population. This study sheds light on this issue and therefore was conducted to verify the hypothesis.

## MATERIALS AND METHODS

### Study area and sampling technique

This web-based cross-sectional study was conducted among five provinces of the Dominican Republic population within a period of June to July, 2020. The data collection instrument was an online administered questionnaire. Five provinces included in the study were described as the Ozama region (comprising Santo Domingo and the National District), Santiago, La Vega and Duarte. The study area was chosen due to the significant mortality trend from COVID-19 [36,37]. Overall, a total of 1195 participants who completed the survey questionnaire were included for the study. Purposive sampling was adopted for participants’ recruitment from the targeted provinces.

### Data collection instrument

The data collection instrument exploited in the study was a short and precise web-based self-designed questionnaire containing closed-ended questions adapted to the target population. The closed-ended items enabled to obtain specific responses from the respondents. The questionnaire comprised four sections: the first section (6 items) obtained sociodemographic data from the respondents, the second section (2 items), third section (1 item) and the fourth section (2 items) were designed to obtain information on the ‘perception of COVID-19 origin’, ‘perception of social distancing’ and ‘knowledge of social distancing’, respectively. An open-ended question about the year of birth was included with the purpose of confirming the age of the participants, due to the easy access that adolescents have to the internet in the Dominican Republic, according to our experience. A pilot study utilising 65 respondents was conducted prior to the actual data collection. To avoid instrumentation which introduces bias to the research, participants’ data for the pilot testing were not selected again for the main study.

### Data collection procedure

A *SurveyMonkey™* collector web link containing the self-designed questionnaire was distributed among participants from the study site at the time of data collection. Individuals aged ≥ 18 years and only from the Dominican Republic were eligible for participating in the study. The survey link was sent as a message via the social media platforms WhatsApp*™*, Facebook*™* and Instagram*™*. The forgoing are the most commonly utilized social media platforms among the Dominican Republic populace [38,39].

### Statistical analysis

Collected data from *SurveyMonkey™* was exported into an excel spreadsheet (*Microsoft Excel 365®, 2016*) for data cleaning, and the dataset was analyzed using SPSS software (IBM SPSS® version 21). Descriptive statistics (mean and standard deviation) were computed to describe the perception and knowledge about the origin of COVID-19 among the study participants, whilst frequencies and percentages were performed to describe the sociodemographic variables (gender, age, province, educational level and monthly income). Stepwise multiple linear regression (MLR) was adopted to estimate the perception of COVID-19 origin using selected sociodemographic variables (gender, age, educational level and income) as predictor variables. Further, one-way multivariate analysis of variance (one-way MANOVA) was performed to determine whether any differences existed between independent groups on more than one continuous dependent variable. The dependent variables were described as ‘perception and knowledge on social distancing’ and the independent variable was ‘perception of COVID-19 origin’. For the purpose of the present study, a *P* value less than 0.05 at 95% confidence interval (P < 0.05) was deemed statistically significant.

## RESULTS

### Descriptive statistics derived from the measurement of participants sociodemographic

A total of 1195 respondents successfully completed the online questionnaire survey. Exactly 597(50.0%) were males whilst the remaining 598(50%) were females. The age (years) of the participants were recorded as categorical values in this order: > 25-44 years (n = 630, 52.2%), > 45-64 years (n = 284, 23.8%), > 18-24 years (n = 211, 17.7%), > 65 years (n = 70, 5.9%). The overall average age was documented as the mean value 2.18 (± 0.786), thus falling in the age range 25-44 years. About 244 (20.4%) of the respondents originated from Santiago, 723(60.5%) from Santo Domingo and the National District, 124 (10.4%) from La Vega and 104(8.7) from Duarte. Regarding education level, 93(7.8%) were in primary school, 247(20.7%) in secondary or elementary school, 95(7.9%) had technical/vocational degree, 611(51.1%) were university students, 101(8.5%) had professional degree, 41(3.4%) had master’s degree and only 7(0.6%) had doctorate (PhD) degree. The household income in Dominican Pesos (DOP) was also explored. 440 (36.8%) of the respondents earned less than RD$ 41,164 (DOP) per month (low income), 46 (3.8%) and 335 (28%) of the respondents earned exactly RD$ 41,164 (DOP) per month (average income) and more than RD$ 41,164 (DOP) per month (high income), respectively. About 217 (18.2%) did not know their monthly income whilst 157(13.1%) decided to keep their monthly income confidential. Distribution of sociodemographic characteristics among the participants is presented in Table 1.

**Table 1.** Descriptive statistics on gender, age, province, education and monthly income among the study participants

| Variables |  | Frequency (n) | Percentage (%) |
| --- | --- | --- | --- |
| Gender | Male | 597 | 50 |
|  | Female | 598 | 50 |
| Age/years | 18-24 | 211 | 17.7 |
|  | 25-44 | 630 | 52.2 |
|  | 45-64 | 284 | 23.8 |
|  | ≥ 65 | 70 | 5.9 |
| **Province** | Santiago | 244 | 20.4 |
|  | Ozama region | 723 | 60.5 |
|  | La Vega | 124 | 10.4 |
|  | Duarte | 104 | 8.7 |
| **Education** | Primary school | 93 | 7.8 |
|  | Secondary school | 247 | 20.7 |
|  | Technical/Vocational degree | 95 | 7.9 |
|  | University student | 611 | 51.1 |
|  | Professional degree | 101 | 8.5 |
|  | Master's degree | 41 | 3.4 |
|  | Doctorate/PhD | 7 | 0.6 |
| **Monthly Income** | Low | 440 | 36.8 |
|  | Average | 46 | 3.8 |
|  | High | 335 | 28 |
|  | Don't know | 217 | 18.2 |
|  | Prefer not to say | 157 | 13.1 |

### Perception and knowledge about the origin of COVID-19

Questions regarding perception about the origin of COVID-19 among the study participants (Table 2) were assigned values on a five-point Likert scale format (1-mostly false, 2-false, 3-neutral, 4-true, 5-mostly true for the perceptions about the origin of COVID-19; 1-strongly disagree, 2-disagree, 3-neutral, 4-agree, 5-strongly agree for the perceptions about social distancing). Since the scales were five-point Likert-type scales format, three (3), the mid-value, was chosen as an average value so that, mean scores below it, were considered a poor response and vice versa. Further, the analyses of the responses were computed in terms of the percentage of the respondents who either “Affirmed” or “Rejected” a given statement. If the summation percentage of respondents who indicated ‘mostly true’ or ‘true’ (or ‘strongly agree’ or ‘agree’) exceeded the percentage of active respondents who revealed ‘mostly false’ or ‘false’ (or ‘strongly disagree’ or ‘disagree’), then the statement was said to have been “Affirmed” in the study and vice versa.

**Table 2.**
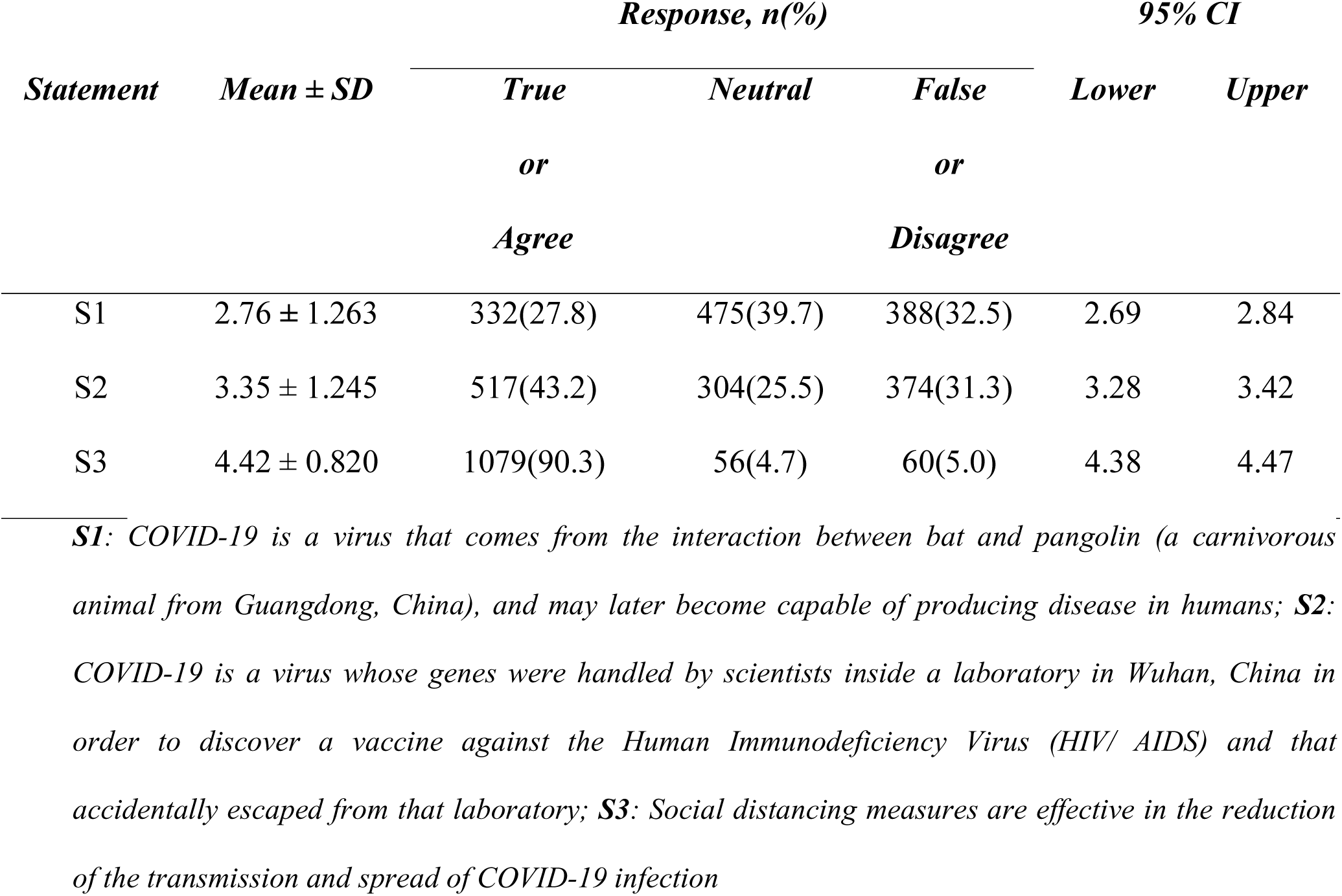
Perception and knowledge about the origin of COVID-19

Our study showed that 332 (27.8%) of the respondents affirmed that ‘COVID-19 is a virus that comes from the interaction between bat and pangolin (a carnivorous animal from Guangdong, China), and may later become capable of producing disease in humans (S1)’. About 517 (43.2%) of the study participants, ‘COVID-19 is a virus whose genes were handled by scientists inside a laboratory in Wuhan, China in order to discover a vaccine against the Human Immunodeficiency Virus (HIV/ AIDS) and that accidentally escaped from that laboratory (S2)’. A greater proportion of the respondents (n = 1079, 90.3%) affirmed that ‘Social distancing measures are effective in the reduction of the transmission and spread of COVID-19 infection’.

### Stepwise Multiple linear regression model for estimating the influence of sociodemographic characteristics on the ‘perception of COVID-19 origin’ and knowledge of social distancing

Multiple linear regression (MLR) equations with corresponding standard error of estimate (SEE) and coefficient of determination (R^2^) were used to estimate the influence of selected sociodemographic characteristics on the ‘perception of COVID-19 origin’ and ‘knowledge of social distancing’ among the respondents (Table 3). No sociodemographic characteristics significantly predicted (*P* > 0.05) individuals’ perception about the origin of COVID-19, except for the level of education (*P* = 0.017). Further, the findings revealed that only age (*P* = 0.032) and education level (*P* < 0.001) statistically significantly predicted ‘knowledge about social distancing’. The coefficient of determination (R^2^) for Perception of COVID-19 origin (R^2^ = 0.009) and Knowledge of social distance (R^2^ = 0.032) revealed that only 0.9% and 3.2% of variations, respectively, may be associated with the sociodemographic background of the participants.

**Table 3.** Stepwise multiple linear regression model for assessing the influence of sociodemographic characteristics on the perception of COVID-19 origin

| Dependent variable | Model | Coefficient (B) | ±SEE | $R^2$ | Sig |
| --- | --- | --- | --- | --- | --- |
| Perception of COVID-19 origin | Constant | 3.623 | 0.184 | 0.009 | .000 |
|  | Gender | 0.028 | 0.073 | 0.000 | .697 |
|  | Age | 0.021 | 0.046 | 0.000 | .655 |
|  | Educational level | -0.070 | 0.029 | 0.006 | .017 |
|  | Income | -0.044 | 0.025 | 0.004 | .078 |
| Knowledge of social distancing | Constant | 0.728 | 0.052 | 0.032 | .000 |
|  | Gender | 0.016 | 0.020 | 0.002 | .436 |
|  | Age | -0.028 | 0.013 | 0.005 | .032 |
|  | Educational level | 0.008 | 0.019 | 0.028 | <.001 |
|  | Income | 0.007 | 0.016 | 0.001 | .597 |

*SEE: Standard error of the estimate*

### One-way MANOVA test for the difference between “knowledge and perception on social distancing” and “perception of COVID 19 origin”

This study further estimated the relationship between the “perception of COVID 19 origin” and “knowledge and perception on social distancing”. The dependent variables were described as “knowledge and perception on social distancing” and the predictor variable or the independent variable was documented as “perception of COVID 19 origin”. A *P* value of less than 0.05 (*P* < 0.05) was considered statistically significant.

Using an alpha level of 0.05 (95% confidence interval), we observe that the MANOVA test produced a statistically significant result; Wilk’s Lamba = 0.967, *F* (4, 1190) = 5.105, *P* <0.001. This significant F indicates that there are significant differences among the “perception of COVID 19 origin” groups on a linear combination of the two dependent variables (knowledge and perception on social distancing). This disclosure explicates that perception of COVID-19 origin were statistically significant (*P* = <0.001) with the measures of the dependent variables (knowledge and perception on social distancing). Distribution of the Multivariate test is presented in Table 4.

**Table 4.**
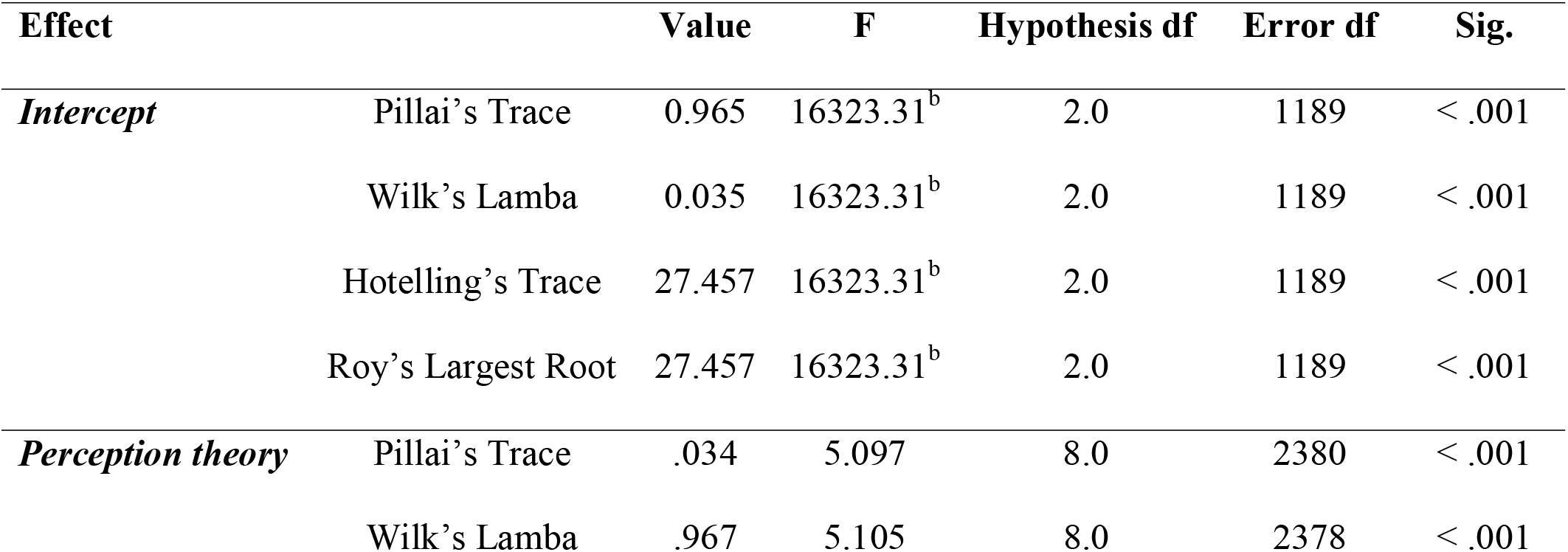
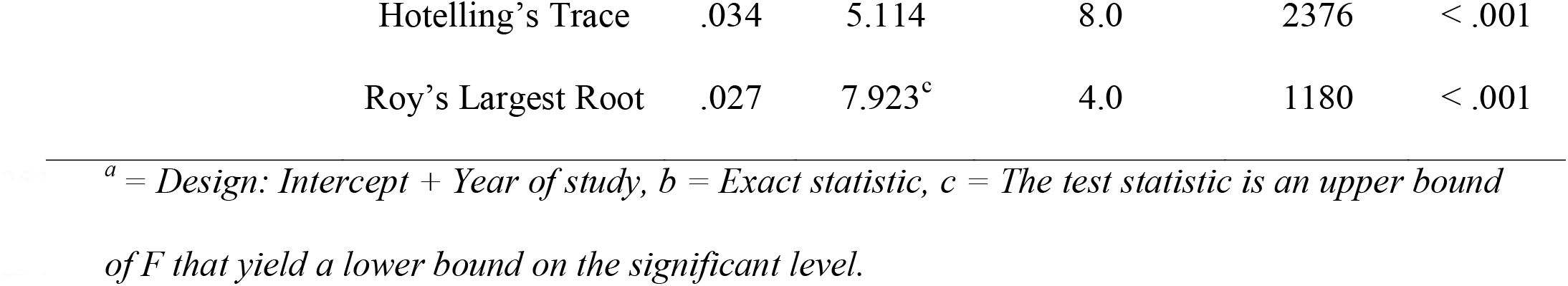
Distribution on multivariate test^a^ (MANOVA test) for the difference between “knowledge and perception on social distancing” and “perception of COVID 19 origin”

Since the MANOVA test was significant, we then examined the univariate ANOVA results to look at the association between the awareness of social distancing and the origin of COVID-19. Follow-up univariate ANOVAs indicated that both knowledge and perception on social distancing were statistically significantly different for participants’ perception about the origin of COVID 19; F (4, 1190) = 5.370, *P* = <0.001, multivariate ŋ^2^ (partial eta squared) = 0.018 and F (4, 1190) = 4.685, *P* = 0.001, multivariate ŋ^2^ (partial eta squared) = 0.016 respectively. The partial eta squared of 0.018 and 0.016 explains that only 1.8% and 1.6% of the multivariate variance in the dependent variable; perception and knowledge of social distancing, respectively, is associated with the group factor (perception of COVID-19 origin). Distribution on the follow-up univariate ANOVAs is presented in Table 5.

**Table 5.** Distribution on the follow-up univariate ANOVAs to test the difference between the dependent variables and the factor group (Perception of COVID 19 origin)

| Source | Dependent Variable | Type III sum of square | df | Mean square | F | Sig. |
| --- | --- | --- | --- | --- | --- | --- |
| Corrected model | PSD | 14.218<sup>a</sup> | 4 | 3.555 | 5.370 | <.000 |
|  | KSD | 2.327<sup>b</sup> | 4 | .582 | 4.685 | .001 |
| Intercepts | PSD | 18477.65 | 1 | 18477.654 | 27915.5 | <.000 |
|  | KSD | 691.94 | 1 | 691.935 | 5572.9 | <.000 |
| **Perception theory** | PSD | 14.218 | 4 | 3.555 | **5.37** | **<.000** |
|  | KSD | 2.327 | 4 | .582 | **4.69** | **.001** |
| Error | PSD | 787.678 | 1190 | .662 |  |  |
|  | KSD | 147.752 | 1190 | .124 |  |  |
| Total | PSD | 24193.0 | 1195 |  |  |  |
|  | KSD | 1019.0 | 1195 |  |  |  |
| Corrected Total | PSD | 801.896 | 1194 |  |  |  |
|  | KSD | 150.079 | 1194 |  |  |  |

$a = R\text{-squared} = 0.018$ ; $b = R\text{-squared} = 0.016$ ; PSD – Perception about social distancing; KSD – Knowledge about social distancing

## DISCUSSION

To date, no research has been conducted in the Dominican Republic focusing on associations between the perceptions about the origin of COVID-19 and the knowledge and perception about social distancing among the general population. Therefore, the present study was conducted to ascertain the associations between the triage (perceptions regarding the origin of COVID-19, knowledge about social distancing and perception about social distancing), as well as the socioeconomic status of the population.

The results of the study indicate that the majority of the participants’ perception is ‘COVID-19 emerged from genetic manipulation by scientists within a laboratory in Wuhan, China, whose purpose was to produce a vaccine against HIV, and spread from an accidental escape’ [16,19-22], while the remaining minority of the individuals’ perception is ‘COVID-19 emerged from the genetic interactions between bat’s and pangolin’s genomic, and may later become capable of producing infection in humans’[8-15].

Most of the participants perceived social distancing as an effective preventative measure for COVID-19 infection and have ample knowledge about it, results that match those of other studies such as that of Taghrir et al.[40]. Since social media outlets have been the primary reported source for research of information regarding COVID-19 by participants in similar studies [34], our findings suggest that the message about social distancing is being propagated correctly via social media, the news and, most importantly, international health organizations [24]. Interestingly, however, this is the same way misinformation about conspiracy and contradicting scientific theories regarding the origin of COVID-19 have also been reproduced [20-22].

A significant association was found between those that perceived the origin of the virus coming from manipulation within a laboratory and their knowledge and perception about social distancing, supporting our hypothesis that there is a relationship between these variables. Although we cannot specifically prove a causal association, these results suggest that the controversy surrounding the origin of COVID-19 and the misinformation disseminated through mass media may also influence other aspects related to individuals’ perceptions and their behavior towards the virus, such as the lack of compliance to government protocols [35,41] and social distancing. These perceptions may also promote distrust in the scientific community, a matter that scientists have tried to raise awareness about [42-47].

Most of the participants in this study were young university students, followed by those with a professional degree. Interestingly, among the sociodemographic characteristics reported, the educational level was the only variable significantly associated with the perceptions about the origin of COVID-19, while both educational level and age were, at the same time, found to be associated with the knowledge about social distancing. However, the variations in the perceptions about the origin of COVID-19 and the knowledge about social distancing within the sample were only 0.9% to 3.2%, respectively, a result that reveals, perhaps, that other sociodemographic factors were not considerably correlated with these variables. These findings contradict those of Bhagavathula [34], where factors such as a professional degree and age of participants were found to be significantly associated with a poor perception and knowledge regarding COVID-19. Further, our findings differ from those in the study of Morinha et. al [35], where the lower the educational level, the more the individuals considered the virus a result of genetic manipulation within a scientific laboratory.

Particularly, the results of our study may suggest that the lack of education level is not the main factor related to perceptions regarding the source of the virus. We think that this phenomenon, resembling the ‘polarization effect’, may support the tendency of individuals with a higher educational level and greater knowledge in science to have more polarized beliefs [48]; a propensity that could be incited by the way the information regarding the origin of COVID-19 is being reproduced in the media. Nonetheless, it is important to highlight that our study differs in several aspects compared to the aforementioned studies, such as the scenario where their research was conducted, and the target populations, and therefore, their results may be not reproducible to our study’s setting. Further, the data collected about these variables may suggest that other sociodemographic factors could be potentially involved with the knowledge and perception about social distancing. Further studies are necessary to be conducted to help clarify these surprising findings.

### Study limitations

A web-based approach was adopted for the purposes of collecting data from the participants. Hence the data collection was made through varied social media platforms which is relatively less utilized among the aged and persons with low socioeconomic background. This approach albeit regarded as an effective and innovative considering the current pandemic situation [49] may introduce a high level of biasness among the group of selected participants for the study. For instance, among the Dominican Republic populace, the average profile of social media users comprises young people from urban areas with a relatively high socioeconomic status compared to the average in other countries of the region [38,39]. Further, about 32.5% of the overall Dominican population do not have access to the internet [50]. This directly implies, this group of the population will be unfairly ruled out automatically. Similar drawbacks have been elaborated by previous researchers [49,51]. Furthermore, another limitation was the difficulty in finding current information about the total population of the country and the national income per capita in public databases; however, for this purpose, data available from the national office of statistics and the central bank of the Dominican Republic [52,53] was used to obtain the most accurate estimation of these values.

Nevertheless, a self-administered online questionnaire is not only an effective and innovative tool at the forefront of the current situation, but provides a relatively feasible monitoring of potential non-respondents, as well as a reduction in the implementation time of the collection instrument, the overall cost of the study [51] and, most importantly, allows compliance with social distancing and other preventive measures imposed by the Dominican government authorities to reduce COVID-19 infection.

## CONCLUSIONS

Our study strongly suggests that more attention needs to be paid to the misinformation regarding the origin of COVID-19 in social media and the news. Further variables are also warrant studying, such as the attitudes of the participants towards social distancing, in order to find associations with these factors and their perceptions about the origin of COVID-19. We acknowledge that inquiring into the perceptions of people from rural areas is also of vital importance and is a matter that demands more research.

## Data Availability

The data supporting the findings of this study are openly available in the data repository figshare. DOI: 10.6084/m9.figshare.12915422.

https://figshare.com/articles/dataset/PercepCovid19OriginSocialdist_SurveyMonkey_Data_xlsx/12915422

## Acknowledgements

We thank all the anonymous participants for providing their responses voluntarily in the survey and Dr. Alondra C. Sepúlveda La Hoz, Marlenis A. Bueno García and Dr. Anabel Castillo Espaillat for their contributions in the promotion and distribution of the survey on social media. We also want to express our gratitude to Dr. María Zunilda Nuñez Payamps, internal medicine specialist and director of the clinical research center Centro de Investigaciones Biomédicas Y Clínicas (CINBIOCLI) and the Dominican Republic’s site centers of the Harvard’s Principles and Practices of Clinical Research (PPCR) for her contributions to the development of ethical and quality research in the Dominican Republic.

## Data availability statement

The data that support the findings of this study are openly available in the data repository figshare at 10.6084/m9.figshare.12915422.

## Supporting information

**S1 fig. Diagram illustrating the process of participants’ recruitment and purposive sampling**. An estimated total of 23,000 participants were reached through different social media platforms. From those, only the results from the responses of 1,195 participants that successfully completed the survey and met the inclusion criteria were analyzed. Note that Ozama region comprises Santo Domingo and the National District provinces that together are also known as the capital city of the Dominican Republic.

## Notes

### Competing Interest Statement

The authors have declared no competing interest.

### Funding Statement

The author(s) received no specific funding for this work.

### Author Declarations

The research protocol was approved by ETIKOS, an ethical committee of the network of institutional review boards of the National Council of Bioethics of the Dominican Republic (CONABIOS) under the registration number CEI-E-2020-04 effective on June 4th, 2020. A written informed consent in electronic format, corresponding to the first pages of the data collection instrument, was sought from the participants prior to recruiting them for the study.

### Summary of Updates

The title of the manuscript was updated from an unofficial short running title to the official full title.

