## Supplemental File 1 for "Misinformation on covid-19 origin and its relationship with perception and knowledge about social distancing: A cross-sectional study"

**Figure 1- Diagram illustrating the process of participants' recruitment and purposive sampling.**

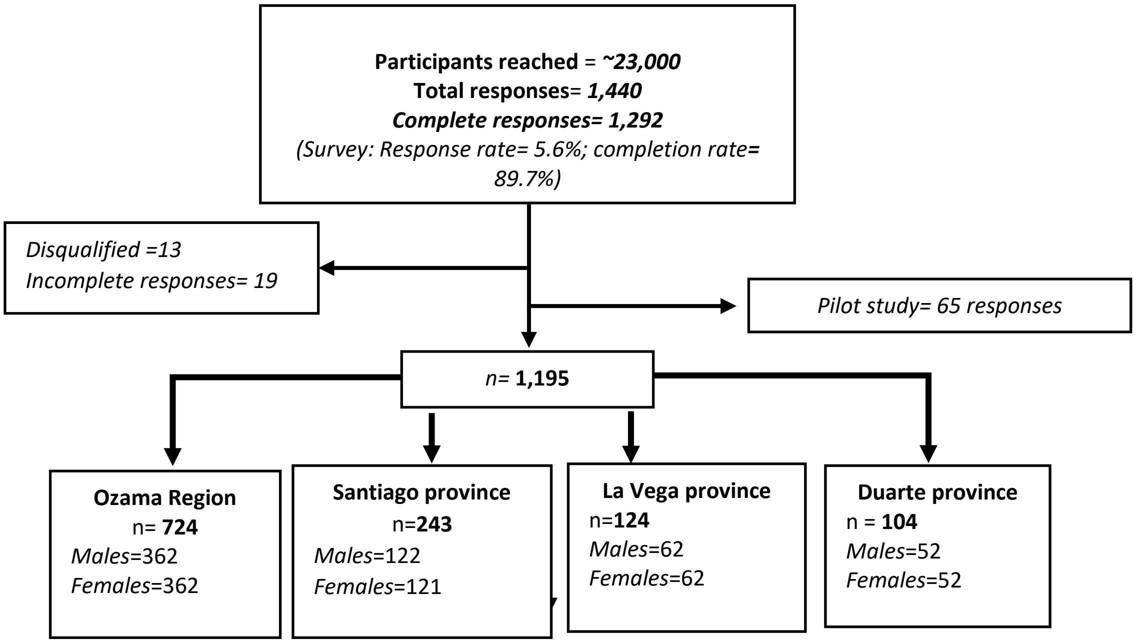


**Figure 1- Diagram illustrating the process of participants' recruitment and purposive sampling.** An estimated total of 23,000 participants were reached through different social media platforms. From those, only the results from the responses of 1,195 participants that successfully completed the survey and met the inclusion criteria were analyzed. Note that Ozama region comprises Santo Domingo and the National District provinces that together are also known as the capital city of the Dominican Republic.
