## Supplemental File 2 for "Misinformation on covid-19 origin and its relationship with perception and knowledge about social distancing: A cross-sectional study"

### Proyecto PercepCOVID

*Encuesta Online*

ANTES DE EMPEZAR, ES NECESARIO QUE LEA DETENIDAMENTE LOS DATOS DE LAS PÁGINAS SIGUIENTES

**cuyo propósito es el de garantizar la calidad y privacidad de los datos suministrados por usted.**


### Proyecto PercepCOVID

*Encuesta Online*


¿Cuál es el propósito de esta encuesta?

**Esta encuesta está destinada a recolectar datos para una investigación independiente cuyo objetivo es conocer la percepción (opinión) que tienen los dominicanos acerca del origen del COVID-19 y del distanciamiento social. Las doctoras en medicina L.Reyes (República Dominicana), Y. Luciano y L. Ortiz. (Estados Unidos) y el especialista en Medicina Interna Pablo J. Reyes R. son quienes están llevando a cabo este proyecto. Esta investigación ha sido aprobada por un comité de bioética debidamente certificado para asegurar el cumplimiento con los lineamientos del Consejo Nacional de Bioética en Salud (CONABIOS), órgano regulador de la investigación en seres humanos de la República Dominicana.**


### Proyecto PercepCOVID

*Encuesta Online*


¿Por qué ha sido invitado a participar en este estudio y qué implicaciones tiene?

**Ha sido invitado a participar por ser de nacionalidad dominicana, tener más de 18 años de edad y ser testigo de la actual**
**pandemia causada por el COVID-19. Su participación es completamente voluntaria. Puede decidir participar o no en el mismo y, en caso de que así sea, puede retirarse en cualquier momento saliendo de esta ventana, y de ninguna manera será cuestionado ni penalizado por su decisión. Ni usted ni los investigadores recibirán compensación monetaria alguna durante el desarrollo de esta investigación. NO se recogerán datos privados tales como su nombre, dirección IP, teléfono, e-mail, entre otros, y los datos suministrados por usted serán utilizados únicamente para propósitos académicos. Los**

**investigadores NO están afiliados al gobierno dominicano NI a organizaciones con fines de lucro NI tampoco reciben financiamiento externo. Participar en esta investigación NO representa ningún riesgo para su salud. Sin embargo, alguna pregunta de la encuesta podría resultarle incómoda.**

### Proyecto PercepCOVID

#### *Encuesta Online*

¿Cuáles son los beneficios que obtendrá al participar en este estudio?

**Se espera que los resultados obtenidos a partir de los datos que usted suministre mediante el llenado de esta encuesta sean de beneficio para la salud de la población dominicana.**

**Si tiene alguna pregunta sobre el proyecto, dudas sobre el consentimiento informado, así como también si quiere obtener acceso a los datos personales y/o resultados generales, los investigadores están a su disposición. Sólo debe enviar un correo electrónico a la siguiente dirección: , y sus dudas y/o peticiones serán atendidas a la brevedad posible.**

**CONSENTIMIENTO ELECTRÓNICO:**

**Hacer click en "SIGUIENTE" indica que:**

**· Ha leído y comprendido la información explicada anteriormente**
**· Está de acuerdo en participar voluntariamente**
**· Tiene o es mayor de 18 años de edad**


### Proyecto PercepCOVID

#### *Encuesta Online*


* 1. Es usted de la República Dominicana?

- ◯ Sí
- ◯ No

* 2. A qué provincia pertenece?

- Santo Domingo
- Santiago de Los Caballeros
- Duarte (San Fco. De Macorís)
- La Vega
- No pertenezco a ninguna de las provincias antes mencionadas

3. **Cuál es su año de nacimiento? (Por ejemplo, 1995)**

### Proyecto PercepCOVID

***Encuesta Online***

* 4. **Cuál es su género?**

- Masculino
- Femenino


### Proyecto PercepCOVID

***Encuesta Online***


5. **What is your age? Por favor escoja un rango:**

- Entre 18-24 años de edad
- Entre 25-44-años de edad
- Entre 45-64 años de edad
- Mayor o igual a 65 años de edad

### Proyecto PercepCOVID

***Encuesta Online***

6. Cuál es su nivel académico?

- Escuela primaria
- Nivel medio o bachillerato
- Grado técnico/vocacional
- Estudiante Universitario
- Grado profesional
- Maestría
- Doctorado ó PhD.
- Prefiero no responder esta pregunta

### Proyecto PercepCOVID

***Encuesta Online***

* 7. ¿Aproximadamente, ¿Cuál es el ingreso económico de su hogar en pesos * dominicanos (DOP)? Por favor, elija un rango:

- Menor a RD$ 41,164 (DOP) per month
- Igual a RD$ 41,164 (DOP) per month
- Major a RD$ 41,164 (DOP) per month
- Desconozco la respuesta ya que soy económicamente dependiente de otra(s) persona(s) para subsistir
- Prefiero no contestar esta pregunta

### Proyecto PercepCOVID

***Encuesta Online***

Seleccione su percepción sobre los siguientes enunciados respecto al COVID-19:

**Elija su opinión respecto a cada teoría enunciada.**

* 8. El COVID-19 es un virus que proviene de la interacción entre el murciélago y el pangolín (animal carnívoro de
Guangdong, China), y que pudo luego volverse capaz de producir enfermedad en el ser humano:

- Mayormente verdadero
- Verdadero
- Neutral (Ni falso ni verdadero)
- Falso
- Mayormente falso

* 9. **El COVID-19 es un virus cuyos genes fueron manipulados por científicos dentro un laboratorio en Wuhan, China para descubrir una vacuna contra el Virus de la Inmunodeficiencia Humana (VIH/SIDA) y que escapó accidentalmente desde dicho laboratorio:**

- Mayormente verdadero
- Verdadero
- Neutral (Ni cierto ni falso)
- Falso
- Mayormente falso

* 10. **Las medidas de distanciamiento social son efectivas para disminuir la propagación y transmisión de la infección por COVID-19:**

- Totalmente de acuerdo
- De acuerdo
- Neutral (Ni de acuerdo ni en desacuerdo)
- En desacuerdo
- Totalmente en desacuerdo

### Proyecto PercepCOVID

***Encuesta Online***

Seleccione la respuesta a las siguientes preguntas respecto al distanciamiento social:

**Seleccione la respuesta que usted crea correcta.**

* 11. El distanciamiento social también se conoce como el distanciamiento físico entre personas:

- Verdadero
- Falso

* 12. ¿Cuál de los siguientes enunciados es correcto de acuerdo a instituciones internacionales de Salud (Por ejemplo, El Centro de Control de Enfermedades -CDC)?

- a) Evitar reuniones en grupo de más de 10 personas.
- b) Usar una mascarilla N95 en lugares públicos.
- c) Mantener una distancia de 6 pies (apróximadamente dos brazos de distancia) de otra persona.
- d) Las medidas de distanciamiento social son más flexibles en niños, ya que éstos tienen un riesgo menor de contraer la infección por COVID-19.
