## Supplemental File 3 for "Misinformation on covid-19 origin and its relationship with perception and knowledge about social distancing: A cross-sectional study"

*TRANSLATED DATA COLLECTION INSTRUMENT*

### THE PERCEPCOVID PROJECT

#### ONLINE SURVEY

#### BEFORE YOU BEGIN, YOU MUST READ THE DATA ON THE FOLLOWING PAGES CAREFULLY

*whose purpose is to guarantee the quality and privacy of the data provided by you*

##### What is the purpose of this survey?

This survey is intended to collect data for an independent investigation whose objective is to know the perception that Dominicans have about the origin of COVID-19 and social distancing. Doctors L. Reyes, Y. Luciano and L. Ortiz. (United States) and the specialist in Internal Medicine Pablo J. Reyes R. are those who are carrying out this project. This research has been approved by a duly certified bioethics committee to ensure compliance with the guidelines of the National Council for Bioethics in Health (CONABIOS), the regulatory body for research on human beings in the Dominican Republic.

##### Why have you been invited to participate in this study and what are the implications?

You have been invited to participate because due to your Dominican nationality, being 18 years old or older and being a witness of the current pandemic COVID-19 pandemic. Your participation is completely voluntary. You can decide to participate or not in the study and, if so, you can withdraw at any time by exiting this window, and in no way will you be questioned or penalized for your decision. Neither you nor the researchers will receive any monetary compensation during the development of this research. NO private data will be collected such as your name, IP address, telephone, e-mail, among others, and the data you provide will be used only for academic purposes. The researchers are NOT affiliated with the Dominican government OR any for-profit organizations nor receive external funding. Participating in this research does NOT pose any risk to your health. However, some questions in the survey may be uncomfortable for you.

**Misinformation on covid-19 origin and its relationship with perception and knowledge about social distancing: A cross-sectional study**

***TRANSLATED DATA COLLECTION INSTRUMENT***

**What is the benefit you will get from participating in this study?**

It is expected that the results obtained from the data that you provide by completing this survey will be of benefit to the health of the Dominican population.

If you have any questions about the project, questions about informed consent, as well as if you want to obtain access to personal data and / or general results, the researchers are at your disposal. You just have to send an email to the following address:, and your doubts and / or requests will be answered as soon as possible.

**ELECTRONIC CONSENT:**

Clicking on "NEXT" you are indicating that:

You have read and understood the information explained above

You agree to participate voluntarily

You are 18 years of age or older

*1-**Are you from the Dominican Republic?**

- Yes
- No

*2-**To which province of Dominican Republic do you belong to?**

- Santo Domingo
- Santiago
- La Vega
- Duarte (San Francisco de Macorís)
- I do not belong to any of the aforementioned provinces

**Misinformation on covid-19 origin and its relationship with perception and knowledge about social distancing: A cross-sectional study**

***TRANSLATED DATA COLLECTION INSTRUMENT***

*3-**What is your year of birth? (For example, 1995)**

*4-**What is your gender?**

- **Male**
- **Female**

*5-**What is your age? Please, choose a range:**

- Between 18-24-year-old
- Between 25-44-year-old
- Between 45-64-year-old
- Greater than or equal to 65 years-old

*6-**What academic level have you reach?**

- Primary School
- Elementary and/or High School
- Technical Degree/ Vocational
- University Student
- Professional Degree
- Master Degree
- Doctorate or Ph. D
- I prefer not to answer this question

*7-**What is your household's income in Dominican Pesos (DOP)? Please, choose a range:**

- Less than RD$ 41,164 (DOP) per month
- Equal to RD$ 41,164 (DOP) per month
- Greater than RD$ 41,164 (DOP) per month
- I do not know the answer since I am economically dependent on another person (s) to survive
- I prefer not to answer this question

**Misinformation on covid-19 origin and its relationship with perception and knowledge about social distancing: A cross-sectional study**

***TRANSLATED DATA COLLECTION INSTRUMENT***

**Select your perception of the following statements regarding COVID-19:** *Choose your opinion on each theory stated.*

*8-**COVID-19 is a virus that comes from the interaction between bat and pangolin (a carnivorous animal from Guangdong, China), and may later become capable of producing disease in humans:**

- Mostly true.
- True.
- Neutral.
- False.
- Mostly False.

*9-**COVID-19 is a virus whose genes were handled by scientists inside a laboratory in Wuhan, China in order to discover a vaccine against the Human Immunodeficiency Virus (HIV/ AIDS) and that accidentally escaped from that laboratory:**

- Mostly true.
- True.
- Neutral.
- False.
- Mostly False.

*10-**Social distancing measures are effective in the reduction of the transmission and spread of COVID-19 infection:**

- Strongly agree.
- Agree.
- Neutral.
- Disagree.
- Strongly Disagree.

*11-**Social distancing is also known as physical distancing between people:**

- True.
- False.

**Misinformation on covid-19 origin and its relationship with perception and knowledge about social distancing: A cross-sectional study**

***TRANSLATED DATA COLLECTION INSTRUMENT***

*12-**Which of the following statements is correct regarding social distancing measures according to the Center for Disease Control and Prevention (CDC)?**

A. Avoiding group meetings of more than 10 people.
B. Wearing a N95 mask in public places.
C. Keeping at least 6 feet (approximately 2 arms away) from another person.
D. Social distancing measures are more flexible for children, as these have a lower risk of contracting COVID-19 infection.
